# Total correlation explanation of toxic metal concentrations and physiological biomarkers amongst NHANES participants

**DOI:** 10.1101/2021.09.30.21264332

**Authors:** James Rooney, Stephan Böse-O’Reilly, Stefan Rakete

## Abstract

**Introduction:** Unravelling the health effects of multiple pollutants presents scientific and computational challenges. CorEx is an unsupervised learning algorithm that can efficiently discover multiple latent factors in highly multivariate datasets. Here, we used the CorEx algorithm to perform a hypothesis free analysis of demographic, biochemical, and toxic metal biomarker data.

**Methods:** Our data included 77 variables from 2,750 adult participants of the National Health and Nutrition Examination Survey (NHANES 2015-2016). We used an implementation of the CorEx algorithm designed to deal with the features of bioinformatic datasets including mixed data-types. Models were fit for a range of possible latent variables and the best fit model was selected as that which resulted in the largest Total Correlation (TC) after adjustment for the number of parameters. Successive layers of CorEx were run to discovered hierarchical data structure.

**Results:** The CorEx algorithm identified 20 variable clusters at the first layer. For the majority clusters, the associations between variables were consistent with known associations – e.g. gender and the hormones, estradiol and testosterone were included in the first cluster; blood organic mercury and blood total mercury were grouped in cluster 4, and cluster 6 included the liver function enzymes ALT, AST and GGT. At the second layer, 3 branches of were identified reflecting hierarchical structure. The first branch included numerous physiological biomarkers and several exogenous biomarkers. The second branch included a number endogenous and exogenous variables previously associated with hypertension, while the third branch included mercury biomarkers and some related endogenous biomarkers.

**Discussion:** We have demonstrated the CorEx algorithm as a useful tool for hypothesis free exploration of a biomedical dataset. This work extends previous implementations of CorEx by allowing mixed data-types to be modelled and the results showed that CorEx detected meaningful hierarchical structure. CorEx may facilitate exploration of novel datasets in future.

## Introduction

Pollution and exposure to toxic chemicals is associated with a wide range of health effects, and disease linked to pollution were estimated to be responsible for 9 million deaths worldwide in 2015^[1]^. Exposure to low doses of toxic metals (and other chemicals) is almost ubiquitous amongst the population. Historically, the effects of toxins on health were studied individually and advances in knowledge have led to progressive controls on chemicals associated with toxic effects. This is perhaps most evident in the case of lead, for which progressively lower ‘safe levels’ were defined across decades until it was recognised by the CDC in 2012 that no safe level exists^[2–4]^. Despite advances in the understanding of the links between chemical exposures and disease at lower exposure levels, our understanding of the health effects of exposure to chemical mixtures has evolved more slowly. This has been due to a number of factors including the previous successes of examining one pollutant at a time, the costs associated with measuring multiple pollutants, and the fact the pollutants are typically highly correlated within individuals, which poses challenges for statistical modelling^[5,6]^. However, interest in assessing and understanding multiple exposures has grown in recent years and a variety of diverse approaches have arisen.

Shared biological pathways and pharmacokinetic factors affecting different toxins might be an unrecognised factor reflected in biomarker correlations. For example lead absorption is competitively inhibited by calcium^[7]^, and, while several toxic metals are in large part excreted via the kidneys (inorganic mercury, lead, cadmium, arsenic), some undergo significant hepatic excretion (manganese, organic mercury, copper), with other routes possible too (for example elemental mercury can be exhaled). Nevertheless some metals retain mostly independent metabolic steps such as the long-term storage of lead in bone, or the respiratory excretion of elemental mercury. Therefore, in an individual with for example mild kidney or liver dysfunction, levels of selected metals could be simultaneously affected while others are not. Thus the tendency for some toxic metal biomarkers to be correlated might in part be explained by shared metabolic pathways. A number of statistical approaches have been developed to deal with such complexity. Sophisticated physiologically based pharmacokinetic models (PBPK), which have a long history of use for single pollutants have been generalised to model multiple exposures^[8]^. however this approach requires detailed scientific understanding of the underlying physiological and pharmacological properties of each chemical of interest^[8]^. On the other hand a number of machine learning based approaches have arisen with BKMR (Bayesian Kernel Machine Regression), in particular, emerging as a popular method for evaluating multiple environmental exposures as risk factors for a given outcome^[5,9]^. BKMR regresses an outcome on a flexible mixture function of proposed environmental pollutants using Bayesian kernel regression and can select from exposure variables using a hierarchical extension, however BKMR is computationally demanding. Another approach is the use of environmental risk scores (ERS)^[6,10]^, a two-step method which may use BKMR (or other machine learning approaches) as the first step to select variables which are then used to compute a risk score, before this in turn is regressed against the outcome of interest.

CorEx is an information theoretic unsupervised learning algorithm that uses total correlation explanation to discover structure in high dimensional data and can efficiently discover multiple latent factors when provided with input variables with high multivariate information^[11–13]^. CorEx can also order those latent factors by the Total Correlation (TC) of the multivariable information of the variables assigned to each latent factor, and successive CorEx runs can be used to discover hierarchical structure^[11–13]^. CorEx has been used to discover biologically interpretable subgroups gene expression groups in ovarian tumour samples^[13]^. Our aim is to apply the CorEx algorithm to NHANES (The National Health and Nutrition Examination Survey) routine biochemical and toxic metal biomarker data in an exploratory analysis to discover hierarchical latent structure within the dataset.

## Methods

### Study Population

The National Health and Nutrition Examination Survey (NHANES) is a long running surveillance project that combines interviews, clinical assessments, clinical measurements and laboratory testing in a representative sample of the US population (https://www.cdc.gov/nchs/nhanes/). It has received ethical approval from the National Center for Health Statistics Ethics Review Board and the participants provided their written informed consent to participate in this study. The study now runs on a continuous basis, although the exact measurements performed vary from year to year. Our analysis includes all adult NHANES participants from the 2015-2016 NHANES data release for whom toxic metals measurements were performed (N = 2,750). Data files including age, BMI and variables relating to blood and urine toxic metals, urinary iodine, and standard haematological assays, biochemical assays including liver and kidney function tests, blood cholesterol and blood glucose were included in the analysis. Data from adults only was included (ages 20 and up) since not all measurements were performed on children.

### Total Correlation Explanation

BioCorEx is an implementation of total correlation explanation designed to model data with characteristics typical of biomedical datasets – i.e. missing data, continuous variables, and severely under-sampled data^[11–14]^. The CorEx algorithm aims to reconstruct latent factors optimised to explain as much of the dependence in the data as possible. Total Correlation (TC), also known as multi-information or multivariate mutual information quantifies dependence amongst a group of variables^[15]^. Given a set of multivariate random variables X ≡ X_1_,…, X_n_ with an associated probability distribution *p*(*X=x*) we can write the marginal probability for a single variable as *p*(*X*_*i*_ = *x*_*i*_), total correlation can be defined both in terms of the Shannon Entropy, *H*, or as a Kullback-Liebler divergence, *D*_*KL*_^[11–13]^:

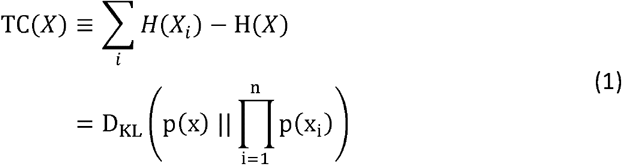

This definition of TC implies that TC is non-negative and can take the value zero only if the variables *X*_1_,…,*X*_*n*_ are independent.

The CorEx algorithm extends the concept of total correlation to facilitate the reconstruction of the set of latent factors *Y = Y*_1_,…, *Y*_*m*_ that minimise the value of TC after condition X on Y^[11–13]^. Thus, we can define the correlation of X explained by Y as the difference between the TC of X and the TC of X given Y as follows:

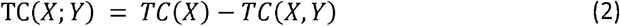

(Note that the semicolon indicates that *TC*(*X*;*Y*) is not symmetric in its arguments – i.e. TC(*X*;*Y*) ≠ TC(*X*;*Y*)). CorEx maximises this expression in a computationally efficient manner given user supplied parameters, m and k, where m defines the number of latent factors *Y* = *Y*_1_, …, *Y*_*m*_, and k defines the number of discrete values each hidden factor can take, or in other words how flexible the Y factors can be. Results from differently parameterised CorEx models can be compared via the resulting value of *TC*(*X*;*Y*), with higher values for *TC*(*X*;*Y*) indicating a greater amount of explained correlation^[11–13]^. The labels output from CorEx can in turn be used as the input to another layer of analysis via CorEx and in this way a deeper hierarchal correlation structure in the data can be discovered (Fig 1). The BioCorEx implementation of the algorithm extends CorEx to accommodate continuous variables, missing data and to include Bayesian smoothing for under-sampled datasets typical of biomedical datasets ^[12–14]^.

**Figure 1.**
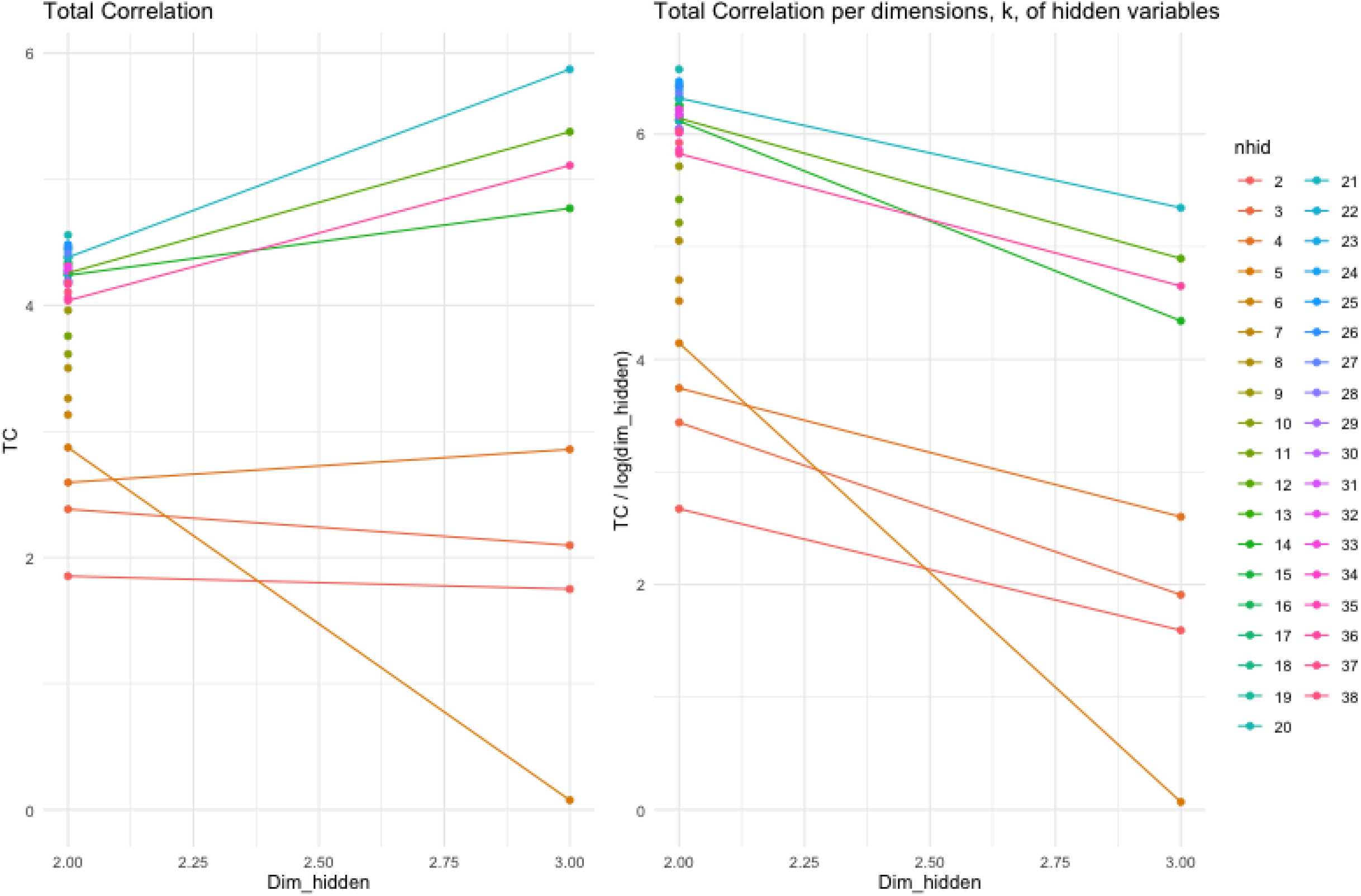
Total correlation and total correlation per k dimensions from CorEx models TC (left) and — (right) vs dim_hidden(k) for CorEx models.

### Statistical Analysis

#### Data preparation

Variables included in the CorEx analysis are shown in Supplementary Table 1. While CorEx can be run on data with missing values, the sampling strategies of the NHANES project meant that some variables are missing for the majority of participants. Therefore we removed any variable with a missing percentage higher than 50%. This resulted in the removal of 3 variables (oral glucose tolerance test, serum insulin concentration and length of fast from food prior to blood glucose measurement). Since, the majority of toxicological and physiological biomarkers are log-normally distributed, we log-transformed all variables except for age, gender and body mass index (BMI).

#### CorEx Analysis

The CorEx model was fit using R Statistical Software 4.0.5^[16]^ using an implementation of BioCorEx written in R (https://github.com/jpkrooney/rcorex)^[17]^ and additional R packages^[18–25]^. This implementation of the CorEx algorithm allows for data to have mixed data types (e.g. Boolean or continuous Gaussian variables), therefore sex and pulse regularity (coded as 0 or 1) were included as Boolean and all others were given the Gaussian marginal description. To mitigate a numerical issue that rarely occurs in a data dependent manner a minimal value of 1 × 10^−200^ was imposed on individual marginal values. The overall marginal was not otherwise limited. In addition, under certain circumstances CorEx can produce negative TC which is undefined, therefore if we detected this circumstance between 10 and 30 iterations of the algorithm the fit was abandoned to avoid redundant computation.

For our analysis we used CorEx to investigate potentially hierarchical data structure in the NHANES variables selected. For layer 1 of the hierarchy we fit CorEx across a range of possible number of hidden clusters up to half the number of included variables (**e**.g. *m* = 1 to 38) and for each value of *m* for *k* = 2 to 3 possible dimensions, giving 76 possible combinations of *m* and *k*. For each of these combinations the CorEx model was fit 25 times and the run which produced the maximal value of TC was retained. Since the maximum TC is partially limited by m and k such that:

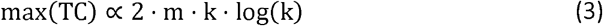

To select the optimal number of hidden clusters, m, and dimensions k, we calculated the value of 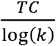 versus k, stratified by m, thus approximately linearizing 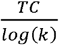 across m and k. Models were then ranked by 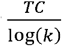, and the maximal model selected as the preferred model. The graph of TC vs iteration for the selected model was then examined. To detect layer 2 of the hierarchical structure this process was repeated this time using the cluster labels from layer 1 as discrete input and only considering a value for *k* = 2. This process was iterated to detect deeper layers of structure until the number of clusters in a given layer = 1. A network graph of the resultant clusters discovered by CorEx was drawn and characteristics of the members of each cluster summarised via descriptive statistics.

#### Analysis code is available from GitHub

https://github.com/jpkrooney/NHANESmetals_corex_Analysis

## Results

Values for 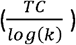 for the 76 combinations of parameters for layer 1 of the CorEx model are shown in Table 1. From ranked results shown in Table 1 and plots of TC vs iterations we selected the CorEx model searching for 20 hidden clusters of dimension 2 as the best-fit model. Figure 1 shows TC and 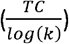 for a selection of *m* and *k* values, while figure 2 shows the TC vs iterations plot for the selected best fit model. While the selected layer 1 CorEx model had a TC of 4.557 nats (natural unit of information), the best layer 2 model had a TC of only 0.391 nats, indicating that layer 2 accounted for some extra correlation in the dataset addition to layer 1. A third hierarchical layer 3 had a TC of just 0.9 × 10^−6^ nats with only one hidden unit indicating very little extra correlation was discovered at the third hierarchical level. Thus, no further layers of structure were investigated. Figure 3 displays a network graph showing the hierarchical structure determined for this CorEx model fit.

**Table 1.** TC / log(k) for *m* hidden clusters with *k* possible dimensions

| Dimensions, k: | 2 | 3 |
| --- | --- | --- |
| Num. of clusters, m |  |  |
| 1 | -0.068 | -2.325 |
| 2 | 1.852 | 1.750 |
| 3 | 2.384 | 2.010 |
| 4 | 2.595 | 2.858 |
| 5 | 2.872 | 0.076 |
| 6 | 3.132 |  |
| 7 | 3.261 |  |
| 8 | 3.502 |  |
| 9 | 3.960 |  |
| 10 | 3.613 |  |
| 11 | 3.760 |  |
| 12 | 4.256 | 5.377 |
| 13 | 4.330 |  |
| 14 | 4.239 | 4.769 |
| 15 | 4.339 |  |
| 16 | 4.384 |  |
| 17 | 4.245 |  |
| 18 | 4.459 |  |
| 19 | 4.374 |  |
| 20 | 4.558 |  |
| 21 | 4.380 | 5.872 |
| 22 | 4.246 |  |
| 23 | 4.433 |  |
| 24 | 4.189 |  |
| 25 | 4.482 |  |
| 26 | 4.454 |  |
| 27 | 4.410 |  |
| 28 | 4.278 |  |
| 29 | 4.193 |  |
| 30 | 4.279 |  |
| 31 | 4.310 |  |
| 32 | 4.272 |  |
| 33 | 4.307 |  |
| 34 | 4.166 |  |
| 35 | 4.061 |  |
| 36 | 4.038 | 5.109 |
| 37 | 4.176 |  |
| 38 | 4.105 |  |

For $m, k$ combinations that are missing TC, no model converged within 200 iterations

**Figure 2.**
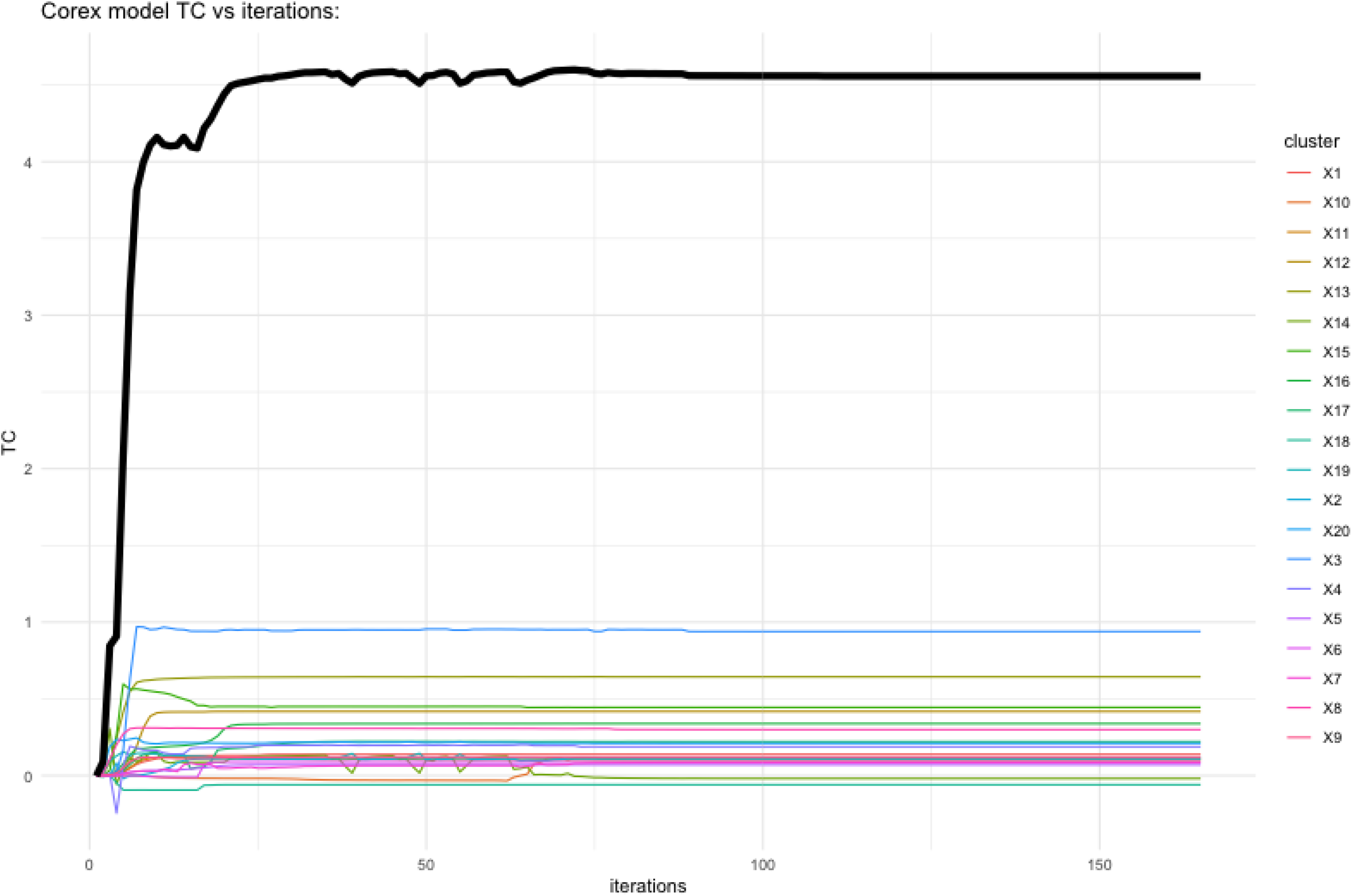
Total correlation vs model iterations for the selected best-fit model.

**Figure 3.**
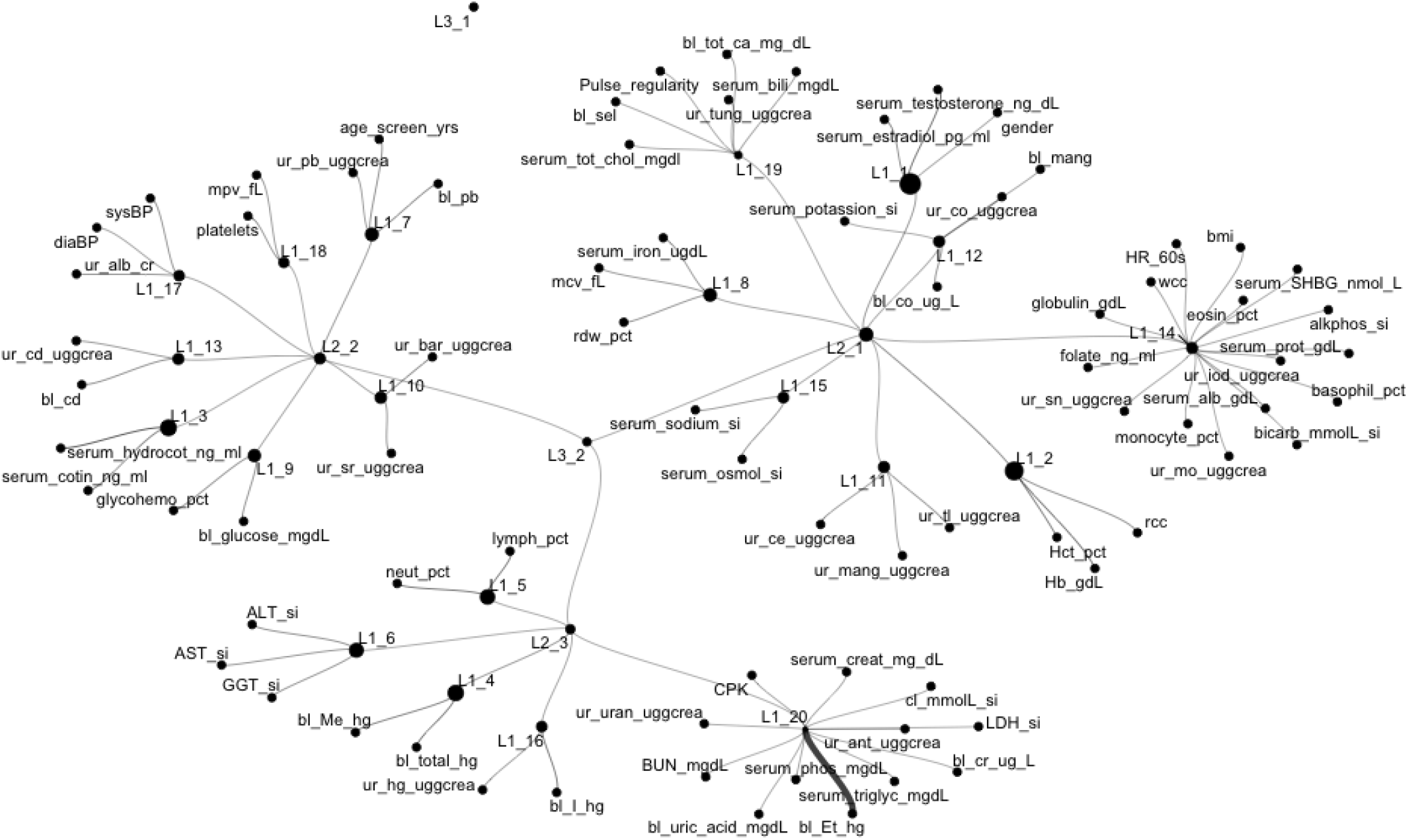
Network graph of NHANES data

Table 2 contains details of the layer1 clusters, including the Total Correlation value for each individual layer 1 cluster with higher TC indicating greater correlation between cluster members. Of the 20 clusters identified, most comprised of a low number of correlated variables. Cluster 1 contains gender and the sex hormones estradiol and testosterone, while cluster 2 contained 3 variables pertaining to red cell physiology. Cluster 3 contained serum cotinine and its metabolite serum hydroxycotinine which are both biomarkers of smoking. Several more small clusters capture toxic metal variables that are closely related: e.g. cluster 4 contains blood organic mercury and blood total mercury, cluster 13 contains blood and urinary cadmium, cluster 16 contains blood inorganic mercury and urinary mercury. Interestingly, cluster 7 contains blood and urine lead concentration and age. Other clusters capture closely related endogenous molecules: e.g. cluster 5 includes two immune function variables – blood lymphocyte and neutrophil percentage, cluster 6 includes 3 markers of liver function (serum AST, ALT and GGT), while cluster 9 contains 2 variables relating to glucose metabolism (blood glucose and glycohemoglobin percentage). Cluster 17 contains systolic and diastolic blood pressure along with urinary albumin to creatinine ratio. Cluster 10 urinary strontium and barium, cluster 11 urinary caesium, manganese, thallium, cluster 12 blood manganese, blood and urinary cobalt and serum potassium.

**Table 2.**
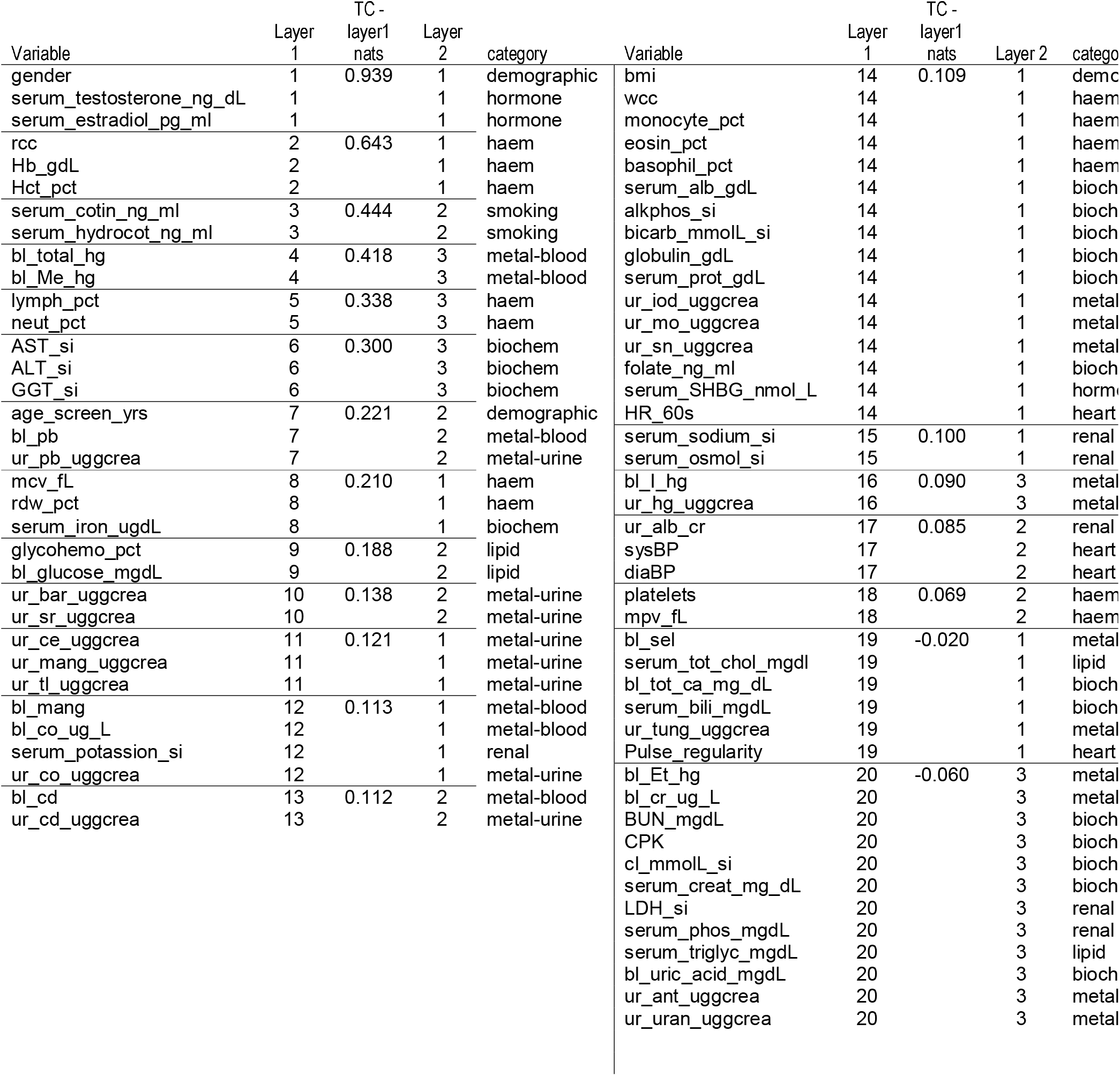
Summary of cluster assignments

Cluster 14 is a large cluster containing 16 variables primarily comprising haematological and biochemical markers, but also containing serum sex-hormone-binding-globulin(SHBG) urinary iodine, molybdenum and tin, as well as heart rate. Cluster 19 and 20 had small negative TC values which we interpret to mean there was no correlation between the variables in these clusters. Cluster 20 included blood ethyl-mercury, urinary antimony and 5 other variables of mixed category, and had a slightly negative TC indicating that these variables were not correlated.

At the second hierarchical level of structure, the first branch had a TC of 0.250 nats and included clusters 1, 2, 8, 11, 12, 14, 15 and 19, therefore including variables mostly relating to endogenous physiology such as gender and sex hormones, routine haematological and biochemical parameters, BMI, manganese and cobalt in blood as well as manganese caesium, thallium, iodine, molybdenum and tin in urine. The second branch at the second hierarchical level had a TC = 0.108 nats, and included clusters 3, 7, 9, 10, 13, 17 and 18, therefore including age and blood glucose and numerous exogenous biomarkers including cotinine and hydroxycotinine, blood and urine lead, blood and urine cadmium, albumin-creatinine ratio and systolic and diastolic blood pressure. Finally, the third branch at the second hierarchical level had a TC = 0.032 nats including clusters 4, 5, 6, 16 and 20. This branch included all the mercury biomarkers, liver function enzymes, lymphocytes and neutrophils and the remaining uncorrelated variables of layer1 cluster 20.

## Discussion

In environmental research the ubiquity of multiple exposures is an ever present challenge. Here we have addressed this challenge through the use of CorEx to explore the data structure of toxic metal and physiological biomarkers. The learned structure in our analysis reveals numerous small variable clusters that include grouped variables with known a-priori biological connections. For example gender and sex hormones (cluster 1), related physiological biomarkers like the liver function makers ALT, AST and GGT (cluster 6), related exogenous chemical biomarkers such as cotinine and hydroxycotinine (cluster 3), or inorganic mercury and urinary mercury (cluster 16). Blood and urine lead biomarkers are clustered together with age, which is an association widely reported in previous cohort studies^[26–28]^. Similarly, cluster 17 groups together systolic and diastolic blood pressure with albumin-creatinine ratio which is again in keeping with previous observations^[29,30]^. Of the smaller clusters, only clusters 10, 11 and 12 produced groupings of variables without explanation readily available from a-priori knowledge.

TC associated with Layer 2 (0.390 nats) was lower than that of layer 1 (4.557 nats). However, the branches of layer 2 also grouped together a mixture of features compatible with known biological factors. The first branch at layer 2 included for the large part routine endogenous physiological biomarkers and some exogenous biomarkers including: urinary caesium, manganese, thallium, cobalt, iodine, molybdenum and tin, as well as blood manganese. The second branch at layer 2 contains the demographic variable age with endogenous biomarkers such as blood glucose and glycol-hemoglobin, platelets and mpv and urinary albumin to creatinine ratio. However this branch also contains a number of exogenous biomarkers including blood and urinary lead and cadmium, cotinine and hydroxycotinine, urinary barium and strontium, systolic and diastolic blood pressure. Interestingly, of the variables included in branch 2 many have been previously associated with blood pressure including cotinine^[31,32]^ (smoking is a well-established risk factor for hypertension^[33–35]^), blood glucose^[36,37]^, lead^[38,39]^, cadmium^[40,41]^, and albumin to creatinine ratio^[29]^. In addition, smoking is source of cadmium and lead exposure in humans^[42–44]^. The third branch at layer 2 included blood total mercury, inorganic mercury and methyl mercury, urinary mercury adjusted for creatinine, lymphocytes and neutrophils and the liver function enzymes AST, ALT and GGT. Correlation of mercury biomarkers is expected since inorganic mercury is calculated from total mercury and methyl-mercury, and mercury biomarkers have previously been associated with lymphocyte and neutrophil counts^[45]^. Furthermore, methyl-mercury undergoes enterohepatic excretion^[46]^ and therefore correlation with liver function is unsurprising.

It is difficult to directly compare our results to that of other machine learning approaches to the NHANES dataset, partly because previous studies included different NHANES variables or used different methodological approaches. For example the Environmental Risk Score (ERS) method has been applied to NHANES environmental exposure variables in several studies and uses a two-stage method to first relate exposure to ERS’s, which are in turn related to outcome(s) of interest^[6,10]^. However these results are difficult to compare to ours due to this two-stage structure. Luo et al. used the BKMR method to examine associations between metal mixtures and markers of kidney function after adjusting for confounders^[47]^. They found that cadmium and lead were associated with both eGFR (calculated from serum creatinine) and albumin-creatinine ratio^[47]^. Interestingly in our study the albumin-creatinine ratio was grouped in the same layer 2 branch as lead and cadmium biomarkers, but serum creatinine was not. However, while methods such as ERS or BKMR apply machine learning within a restricted range of hypotheses determined by pre-defined outcomes, risk factors and adjustment variables, a major strength of CorEx is that it is a fully unsupervised method (as applied here). Additionally, our implementation of CorEx extends on previous versions to allow the inclusion of data of mixed types, that is both Boolean and continuous variables.

## Conclusions

We have demonstrated the application of Total Correlation Explanation to epidemiological data with mixed data types using the CorEx algorithm. The results we obtained after fitting CorEx in a hypothesis-free manner to a typical biostatistical dataset demonstrated that the CorEx algorithm could detect structure consistent with previously established biological relationships typical of the demographic, endogenous and exogenous biomarkers included. This work extends previous implementations of CorEx by allowing mixed data-types to be modelled, and therefore has potential to facilitate exploration of novel datasets in future.

## Supporting information

Strobe checklist

## Data Availability

NHANES data is available from the NHANES website: https://wwwn.cdc.gov/nchs/nhanes/Default.aspx
This manuscript analysis code is available here: https://github.com/jpkrooney/NHANESmetals_corex_Analysis

https://wwwn.cdc.gov/nchs/nhanes/Default.aspx

https://github.com/jpkrooney/NHANESmetals_corex_Analysis

## Acknowledgements

We would like to acknowledge the assistance of Dr Greg Ver Steeg, Information Sciences Institute, University of Southern California for explanations of the Python implementation of CorEx.

