## Supplementary Table 1 for "Total correlation explanation of toxic metal concentrations and physiological biomarkers amongst NHANES participants"

Supplementary Data

Supplementary Table 1. List of variables analysed via CorEx

| NHANES Variable names | Category | Renamed to | Note |  | NHANES Variable names | Category | Renamed to | Note |
| --- | --- | --- | --- | --- | --- | --- | --- | --- |
| RIAGENDR | demographic | gender |  |  | LBXIN | biochemistry | insulin_uUmL |  |
| RIDAGEYR | demographic | age_screen_yrs |  |  | PHAFSTMN | biochemistry | foodfast_mins |  |
| BMXBMI | demographic | bmi |  |  | LBXGLT | biochemistry | ogtt2hr_mg_dl |  |
| LBXWBCSI | haematological | wcc |  |  | LBXSAL | biochemistry | serum_alb_gdL |  |
| LBXLYPCT | haematological | lymph_pct |  |  | LBXSAPSI | biochemistry | alkphos_si |  |
| LBXMOPCT | haematological | monocyte_pct |  |  | LBXSASSI | biochemistry | AST_si |  |
| LBXNEPCT | haematological | neut_pct |  |  | LBXSATSI | biochemistry | ALT_si |  |
| LBXEOPCT | haematological | eosin_pct |  |  | LBXSBU | biochemistry | BUN_mgdL |  |
| LBXBAPCT | haematological | basophil_pct |  |  | LBXSC3SI | biochemistry | bicarb_mmolL_si |  |
| LBXRBCSI | haematological | rcc |  |  | LBXSCA | biochemistry | bl_tot_ca_mg_dL |  |
| LBXHGB | haematological | Hb_gdL |  |  | LBXSCK | biochemistry | CPK |  |
| LBXHCT.x | haematological | Hct_pct |  |  | LBXSCLSI | biochemistry | cl_mmolL_si |  |
| LBXMCVSI | haematological | mcv_fL |  |  | LBXSCR | biochemistry | serum_creat_mg_dL |  |
| LBXRDW | haematological | rdw_pct |  |  | LBXSGB | biochemistry | globulin_gdL |  |
| LBXPLTSI | haematological | platelets |  |  | LBXSGTSI | biochemistry | GGT_si |  |
| LBXMPSI | haematological | mpv_fL |  |  | LBXSIR | biochemistry | serum_iron_ugdL |  |
| URXUMS | renal | ur_alb_mgL |  |  | LBXSTB | biochemistry | serum_bili_mgdL |  |
| LBXSKSI | renal | serum_potassion_si |  |  | LBXSTP | biochemistry | serum_prot_gdL |  |
| LBXSLDSI | renal | LDH_si |  |  | LBXSUA | biochemistry | bl_uric_acid_mgdL |  |
| LBXSNASI | renal | serum_sodium_si |  |  | LBDRFO | biochemistry | folate_ng_ml |  |
| LBXSOSSI | renal | serum_osmol_si |  |  |  |  |  |  |
| LBXSPH | renal | serum_phos_mgdL |  |  |  |  |  |  |
| LBXBPB | metal-blood | bl_pb |  |  |  | metal-urine | ur_iod_uggcrea |  |
| LBXBCD | metal-blood | bl_cd |  |  |  | metal-urine | ur_hg_uggcrea |  |
| LBXTHG | metal-blood | bl_total_hg |  |  |  | metal-urine | ur_bar_uggcrea |  |
| LBXBSE | metal-blood | bl_sel |  |  |  | metal-urine | ur_cd_uggcrea |  |
| LBXBMN | metal-blood | bl_mang |  |  |  | metal-urine | ur_co_uggcrea |  |
| LBXIHG | metal-blood | bl_I_hg |  |  |  | metal-urine | ur_ce_uggcrea |  |
| LBXBGE | metal-blood | bl_Et_hg |  |  |  | metal-urine | ur_mo_uggcrea |  |
| LBXBGM | metal-blood | bl_Me_hg |  |  |  | metal-urine | ur_mang_uggcrea |  |
| LBXBCR | metal-blood | bl_cr_ug_L |  |  |  | metal-urine | ur_pb_uggcrea |  |
| LBXBCO | metal-blood | bl_co_ug_L |  |  |  | metal-urine | ur_ant_uggcrea |  |
| LBXTC | lipid | serum_tot_chol_mgdl |  |  |  | metal-urine | ur_sn_uggcrea |  |
| LBXGH | lipid | glycohemo_pct |  |  |  | metal-urine | ur_sr_uggcrea |  |
| LBXSGL | lipid | bl_glucose_mgdL |  |  |  | metal-urine | ur_tl_uggcrea |  |
| LBXSTR | lipid | serum_triglyc_mgdL |  |  |  | metal-urine | ur_tung_uggcrea |  |
| LBXTST | hormone | serum_testosterone_ng_dL |  |  |  | metal-urine | ur_uran_uggcrea |  |
| LBXEST | hormone | serum_estradiol_pg_ml |  |  | LBXCOT | smoking | serum_cotin_ng_ml |  |
| LBXSHBG | hormone | serum_SHBG_nmol_L |  |  | LBXHCT.y | smoking | serum_hydrocot_ng_ml |  |
